# A NOVEL ELECTROCARDIOGRAPHIC PREDICTION MODEL FOR DIAGNOSING CONCEALED BRUGADA SYNDROME: RESULTS FROM A CLINICAL TRIAL OF AJMALINE PROVOCATION IN HEALTHY SUBJECTS

**DOI:** 10.1101/2023.06.26.23291923

**Authors:** Bode Ensam, Chiara Scrocco, David Johnson, Yanushi D. Wijeyeratne, Rachel Bastiaenen, Belinda Gray, Chris Miles, Yael Ben-Haim, Stathis Papatheodorou, Sanjay Sharma, Michael Papadakis, Brian Devine, Peter W. Macfarlane, Elijah R. Behr

## Abstract

**Background:** Clinical, electrocardiographic, and genomic factors have been associated with the drug-induced type 1 Brugada pattern (DI-T1BP), in response to sodium channel blocker provocation (SCBP). However, prior analyses have been concerned with prediction of the DI-T1BP rather than the validity of the diagnosis of concealed Brugada syndrome (BrS). We sought to analyse and compare the ECG response to SCBP with ajmaline in a cohort of healthy controls (HC) and a definite BrS group (Def-BrS) to develop a diagnostic score.

**Methods:** Healthy controls (HC) were systematically recruited as part of a clinical trial. Following comprehensive cardiovascular screening, eligible subjects underwent SCBP with ajmaline. We identified a Def-BrS cohort, defined as a DI-T1BP and a Shanghai Score (SS) ≥3.5, from consecutive patients with suspected BrS undergoing SCBP with ajmaline using the identical protocol. Def-BrS and HC were divided equally into discovery and validation cohorts. Digital ECG acquisition facilitated automated measurement of ECG parameters. A multivariable analysis compared ECG parameters between the HC and Def-BrS cohorts. A logistic regression analysis identified ECG characteristics that accurately predicted the diagnosis of Def-BrS. This model was then assessed in the validation cohort.

**Results:** Two-hundred-and-forty-eight volunteers completed an online questionnaire, 103 accepted an invitation to undergo further screening and 100 were recruited into the HC group. Three HCs developed a DI-T1BP. From 1241 patients undergoing SCBP, 166 were Def-BrS. There were no demographic differences between the HC discovery and validation groups or between the Def-BrS discovery and validation groups. Following multivariable logistic regression analysis, QRS duration, mean anterior lead ST segment amplitude at baseline, maximum change in QRS duration, anterior ST segment amplitude and QRS area after SCBP, were independently associated with Def-BrS. The combined model was an excellent discriminator for Def-BrS, with an area under the curve of 0.95 [95% confidence interval (CI) = 0.912 – 0.989], P<0.001 in the discovery groups and 0.97 [95% CI = 0.948 – 0.998], P<0.001 in the validation groups.

**Conclusion:** The yield of the DI-T1BP in HCs is 3%. However, there are distinct ECG parameters at baseline and in response to SCBP that favour a definite diagnosis of BrS. These observations permit the quantifiable refinement of the ECG diagnosis of concealed BrS, avoiding the pitfalls of relying upon the DI-T1BP alone.

## INTRODUCTION

The yield of the drug-induced type 1 Brugada pattern (DI-T1BP), across a spectrum of clinical and experimental scenarios has been described previously ^(1-4).^ Additionally, the clinical, electrocardiographic, and genomic factors associated with the development of the DI-T1BP in response to sodium channel blocker provocation (SCBP) have been investigated ^(5)^. However, these prior analyses have been concerned with the prediction of the DI-T1BP rather than the strength or validity of the diagnosis of Brugada syndrome (BrS).

In response to a perceived higher than expected yield of the DI-T1BP in the clinical setting and an uncertainty regarding false positives, an international consensus document downgraded the response to SCBP from diagnostic to probabilistic. With the proposed Shanghai scoring system (SS), an isolated DI-T1BP is insufficient to make a diagnosis of “definite Brugada Syndrome ^(6)^.” The scoring system is, however, based largely on expert opinion.

Furthermore, the response to SCBP is not binary, rather, several temporal ECG changes may be observed, and it is postulated to exacerbate a pre-existent impaired right ventricular outflow tract (RVOT) conduction reserve to unveil the DI-T1BP ^(7)^. The significance of baseline and dynamic ECG traits in affected and unaffected subjects is poorly understood but may provide a quantitative element to the drug-induced BrS phenotype.

We sought to analyse and compare the ECG response to SCBP with ajmaline in a cohort of healthy subjects, Healthy Controls (HC), and clinical patients with a DI-T1BP, thus proposing a quantitative ECG model for the diagnosis of definite concealed BrS.

## METHODS

### Recruitment

#### Healthy Controls (HC)

Subjects responded to online advertisements seeking healthy volunteers as part of an interventional clinical trial, (*ClinicalTrials.gov Identifier: NCT02933437, EudraCT Number 2016-004277-41*), with ethical approval from London -Southeast Research Ethics Committee, (*16/LO/2173*).

A comprehensive online medical questionnaire was used to identify eligible subjects (supplementary data I). Those fulfilling the initial inclusion criteria (supplementary data II), were invited to attend for further eligibility testing with a focused cardiovascular examination, baseline resting 12 lead and high right precordial lead (HRPL) ECG with additional leads placed in the V1 and V2 position in the 2^nd^ and 3^rd^ intercostal spaces, (supplementary data III). A baseline transthoracic echocardiogram (TTE) was also performed. Those with any ECG or structural abnormality outside of normal physiological range were excluded (supplementary data II). Participants were compensated for their time.

#### Clinical cohort

From an existing clinical cohort of consecutive patients investigated at our centre for suspected BrS between 2010 – 2022, we identified those with DI-T1BP and a definite diagnosis of BrS according to a SS ≥3.5 (Def-BrS group). Patients within the clinical cohort were evaluated in a dedicated clinic by experienced clinicians following international recommendations and provided informed consent, (NHS Research Ethics Committee reference 10/H0803/121). The indication for SCBP included unexplained cardiac arrest (UCA) in the subject, or BrS, sudden arrhythmic death (SADS) or UCA in a first-degree relative.

#### Study cohort

The HC and Def-BrS group were equally divided into discovery (HC-Dis and Def-BrS-Dis) and validation groups (HC-Val and Def-BrS-Val) groups respectively.

#### Sodium channel blocker provocation

All subjects underwent SCBP with ajmaline according to a standardised protocol of 1mg/kg (maximum dose 100mg) administered intravenously over 5 minutes. The SCBP test was terminated early once the DI-T1BP was observed, if QRS prolongation increased ≥150% of the baseline measurement, or if a ventricular arrhythmia was encountered. The DI-T1BP was defined as J point elevation ≥0.2mV with coved ST elevation in a right precordial chest lead.

#### ECG acquisition and analysis

SCBP was undertaken using HRPL placement. A continuous ECG was acquired digitally using a CardioSoft resting digital recorder (GE Healthcare, Milwaukee, WI, USA) with a sampling rate of 500 samples/sec and 4.88μV amplitude resolution. The low pass filter was set at 150 Hz. From the continuous ECG recording a 10-second HRPL ECG was obtained at 1-minute intervals from the beginning of drug administration for a minimum of 10 minutes.

ECGs were stored in XML format and imported onto a secure ECG server for analysis. Relevant and novel ECG parameters were measured using bespoke automated ECG analysis software developed in partnership with the Electrocardiology Group, School of Health and Wellbeing, University of Glasgow. Beats with excessive noise were excluded from analysis and a combination of lead-specific and global ECG parameters were analysed using single beats and a derived representative beat respectively.

Quantitative ECG parameters were measured at baseline (time 00:00 of ajmaline administration) and at peak drug effect, (denoted as maximum QRS duration (ms) from a representative beat), which were expressed as delta values (Δ= peak – baseline). See Table 1.0 and supplementary material IV for ECG parameters.

**Table 1.0.** ECG parameters – abbreviations and units of measure.

| Lead Measures | Abbreviation | Units |
| --- | --- | --- |
| **Global Representative Beat Data** |  |  |
| QRS Duration | gQRSDur | ms |
| T wave Duration | gTDur | ms |
| PR Interval | gPR | ms |
| QT Interval | gQT | ms |
| P wave Duration | gPDur | ms |
| **+Precordial Lead Data** |  |  |
| QRS Duration | QRS_Dur | ms |
| T peak to T end interval | Tpe_Int | ms |
| T wave Duration | T_Dur | ms |
| Anterior ST amplitude | antSTJ_Amp | µV |
| §QRS Area | QRS_Area | mV/ms |
| §T wave area | TW_Area | mV/ms |
| §QRST Area (QRS onset to T wave termination) | QRST_Area | mV/ms |
| §ST-J point to Tpeak Area (QRS termination to T wave peak) | STj-Tpeak_Area | mV/ms |
| §Tpeak to Tend Area (T wave peak to T wave termination) | Tpe_Tend_Area | mV/ms |
| **Additional Lead Data** |  |  |
| S wave duration in lead I | S_Dur | ms |
| S wave amplitude in lead I | S_Amp | µV |
| + Precordial lead data = sum of precordial leads / no. precordial leads<br>§ Net area<br>ms = milliseconds, uV = microvolts, mV/ms = millivolts per millisecond |  |  |

#### Shanghai Score

The clinical, familial, and genetic components of the SS were applied according to the recommended scoring system^(6)^. However, to investigate the significance of a baseline type 2/3 Brugada pattern (T2/3BP) as a categorical covariate in the prediction of Def-BrS, an ECG score of 2.0 was given to all who developed the T1BP irrespective of the presence of T2/3BP on the baseline ECG. If a spontaneous or fever induced T1BP was observed at any point during follow up the ECG score was adjusted accordingly.

## STATISTICAL ANALYSIS

### General Statistical Methods

Data were analysed with IBM ® SPSS ® Statistics version 29. Categorical variables are presented as frequencies (percentages) and were compared using the 𝜒^2^ or Fishers exact test where appropriate. For continuous variables, the Kolmogorov–Smirnov test was used to test the distribution of data. The Mann-Whitney test was performed to compare differences between non-normally distributed continuous variables, which are reported as median [1^st^ quartile – 3^rd^ quartile]. Normally distributed continuous variables were analysed using an independent sample T-test and are reported as mean ± standard deviation (SD). A value of P< 0.05 was considered significant in all cases. A paired intragroup analysis of ECG characteristics was undertaken and is provided in supplementary data V.

### Variable Filters and Model Development

Candidate variables from the univariate analysis between the HC-Dis and Def-BrS-Dis groups with a statistical significance of P<0.05 were included in a multivariate binomial logistic regression model.

Following this, variables that were independently associated with the diagnosis of Def-BrS (odds ratio (OR) >1.0 and P<0.05), were included in the predictive model.

To assess for overfitting, we used the Akaike Information Criterion (AIC) and Bayesian Information Criterion (BIC), and we assessed the performance of the proposed model using the validation groups.

## RESULTS

### Demographic and clinical characteristics

Two hundred-and-forty-eight (248) prospective healthy subjects completed the online medical questionnaire, 68 applicants were excluded at this stage due to prior cardiac symptoms or suspicious family history. One-hundred-and-eighty were deemed suitable and were invited for further eligibility testing with 103 attending. Two subjects were excluded because of baseline ECG abnormalities and one subject was excluded due to a borderline TTE abnormality. We did not observe a spontaneous T1BP or T2/3BP in any of the subjects undergoing screening. The final cohort of 100 healthy controls underwent SCBP (Figure 1.0).

**Figure 1.0.**
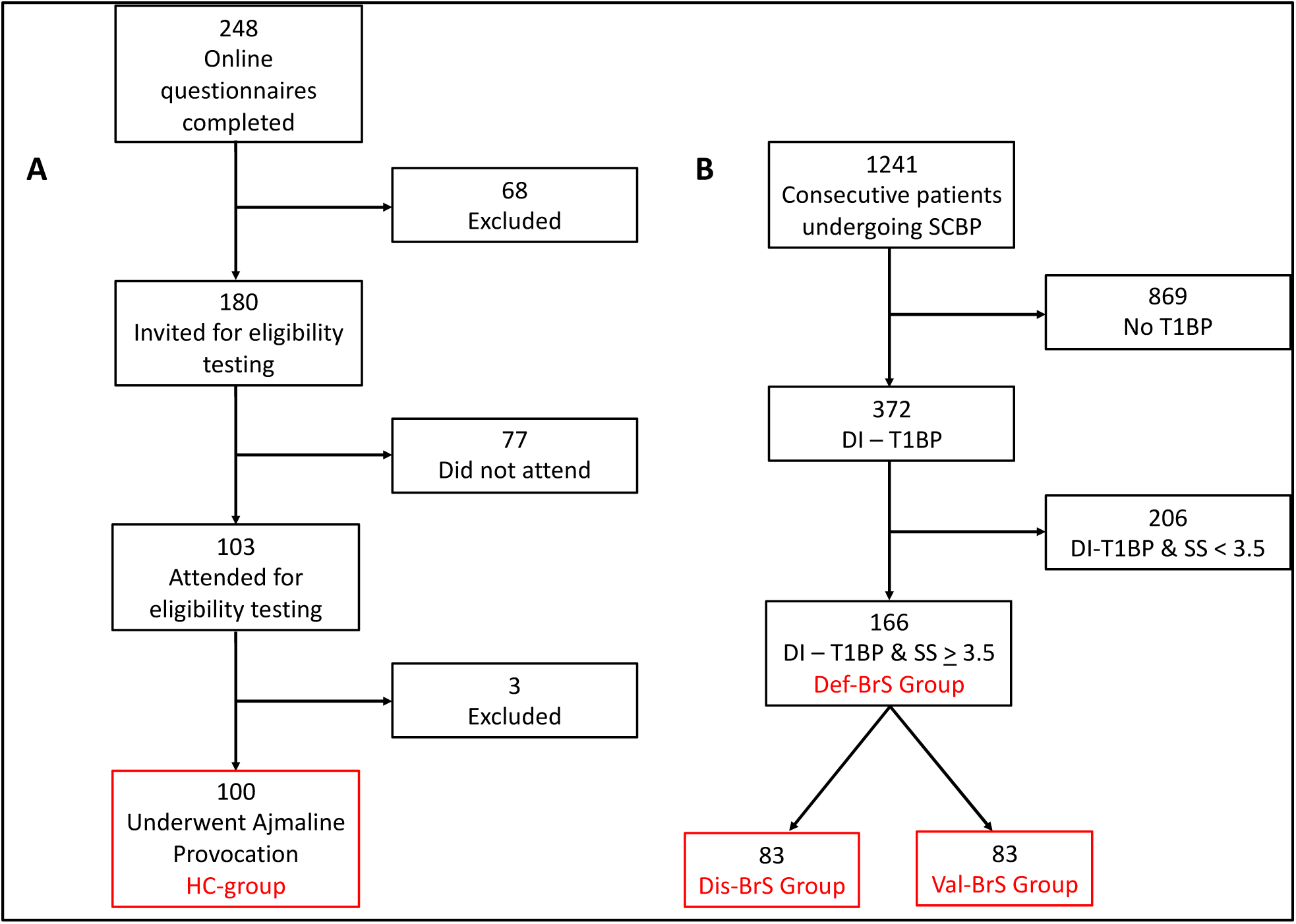
Cohort recruitment flow diagram. A – Healthy controls, B – Clinical cohort. SCBP = Sodium Channel Provocation, HC = healthy controls, Def-BrS = definite Brugada syndrome, Def-BrS-Dis = definite Brugada syndrome discovery, Def-BrS-Val = definite Brugada syndrome validation, HC-Dis = healthy control discovery group, HC-Val = healthy control validation group.

From a clinical cohort of 1241 consecutive patients undergoing AP, 372 (30%) had a DI-T1BP. One hundred-and-sixty-six of these patients, 45% (166/372), had a SS of ≥3.5 and comprised the Def-BrS cohort (Figure 1.0).

Table 2.0 highlights the demographic and clinical characteristics of the overall HC and Def-BrS cohorts. Of note the HC cohort were significantly younger in age than the Def-BrS cohort, median age 24.53 years (Q1 21.78 – Q3 29.04) vs. 42.50 (Q1 28.00 – Q3 59.21), P<0.01. Within the Def-BrS cohort, 31% (51/166) were BrS probands and 33% (55/166) had a history of syncope prior to evaluation. During follow-up 12% (20/166) were found to have a spontaneous T1BP on one or more occasion and 4% (6/166) had spontaneous T1BP associated with fever. The presence of a T1BP in a 1^st^ or 2^nd^ degree relative was the most frequent indication for SCBP in the Def-BrS cohort, 60% (99/166). In the combined Def-BrS group, 10% (17/166) had a baseline type 2/3 Brugada pattern (T2/3BP). However, no HC’s had a baseline T2/3BP (P= 0.02). A pathogenic variant in the *SCN5A* gene was identified in 19% (31/166) Def-BrS subjects. The mean time to peak drug effect was significantly earlier in the Def-BrS group compared to HC group 05min18secs (±2min05secs) vs. 05min59secs (±01min20secs), P=0.01. Mean final SS in the Def-BrS group was 4.3 (±0.75) vs. 0.06 (±0.34) in the HC group.

**Table 2.0.** Overall cohort demographic and clinical characteristics. HC=Healthy Controls. Def-BrS=Definite Brugada Syndrome.

|  | **HC**<br>**N = 100** | **Def-BrS**<br>**N = 166** | **P value** |
| --- | --- | --- | --- |
|  | **Demographics** |  |  |
| **Median Age (years) (Q1 - Q3)** | 24.53<br>(21.78 – 29.04) | 42.50<br>(28.00 – 59.21) | <0.01 |
| **Male Gender** | 52 (52%) | 72 (43%) | 0.14 |
| **Caucasian Ethnicity** | 100 (100%) | 133 (80%) | <0.01 |
| **BrS Proband** | - | 51 (31%) | <0.01 |
| **Syncope** | - | 55 (33%) |  |
| **Baseline T2/3 Brugada pattern** | 0 (0%) | 17 (10%) | 0.02 |
|  | **Ajmaline Indication** |  |  |
| **Healthy Control** | 100 (100%) | 0 (0%) | - |
| **SADS/UCA relative** | - | 39 (24%) |  |
| **T1BP in a relative** | - | 99 (60%) |  |
| **UCA survivor** | - | 11 (7%) |  |
|  | **Family History Score** |  |  |
| **0** | 100 (100%) | 30 (18%) | - |
| **0.5** | - | 12 (7%) |  |
| **1** | - | 17 (10%) |  |
| **2** | - | 107 (65%) |  |
| **Mean (SD)** | 0 | 1.44 (0.81) |  |
|  | **Clinical History Score** |  |  |
| **0** | 100 (100%) | 109 (66%) | - |
| **0.5** | - | 2 (1%) |  |
| **1** | - | 28 (17%) |  |
| **2** | - | 10 (6%) |  |
| **3** | - | 17 (10%) |  |
| **Mean (SD)** | 0 | 0.60 (0.98) |  |
|  | **ECG Score** |  |  |
| **0** | 97 (97%) | 0 (0%) | - |
| **2** | 3 (3%) | 140 (84%) |  |
| **3** | - | 6 (4%) |  |
| **3.5** | - | 20 (12%) |  |
| **Mean (SD)** | 0.06 (0.34) | 2.22 (0.51) |  |
|  | **Genetic Score** |  |  |
| **0.5** | 0 (0%) | 31 (19%) | - |
| **Mean (SD)** | 0 | 0.09 (0.20) |  |
|  | **Ajmaline Provocation** |  |  |
| **Pre - ajmaline SS (SD)** | 0 | 2.1 (0.80) | <0.01 |
| **Final SS (SD)** | 0.06 (0.34) | 4.3 (0.75) | <0.01 |
| **Time to Peak Drug Effect (SD)** | 5m59s (1m20s) | 5m18s (2m05s) | 0.01 |
| **Final dose:Target dose** | 1.00 (0.00) | 0.78 (0.22) | <0.01 |

### Electrocardiographic Characteristics and Analyses

Overall, 3 subjects in the HC group developed the DI-T1BP during SCBP, 3/100 (3%) (Figure 2.0). At baseline, all 3 subjects had a partial right bundle branch block pattern (RBBB) with a sharp r’ but without resting J-point elevation in at least one HRPL. In 2 of these 3 subjects, the DI-T1BP was observed in a single high-right precordial chest lead (HRPL) only, whilst in the remaining subject (subject 3), the DI-T1BP was seen in the 4^th^ and 3^rd^ intercostal spaces (ICS) and was borderline (<0.2mV J-point elevation) in the 2^nd^ ICS.

**Figure 2.0.**
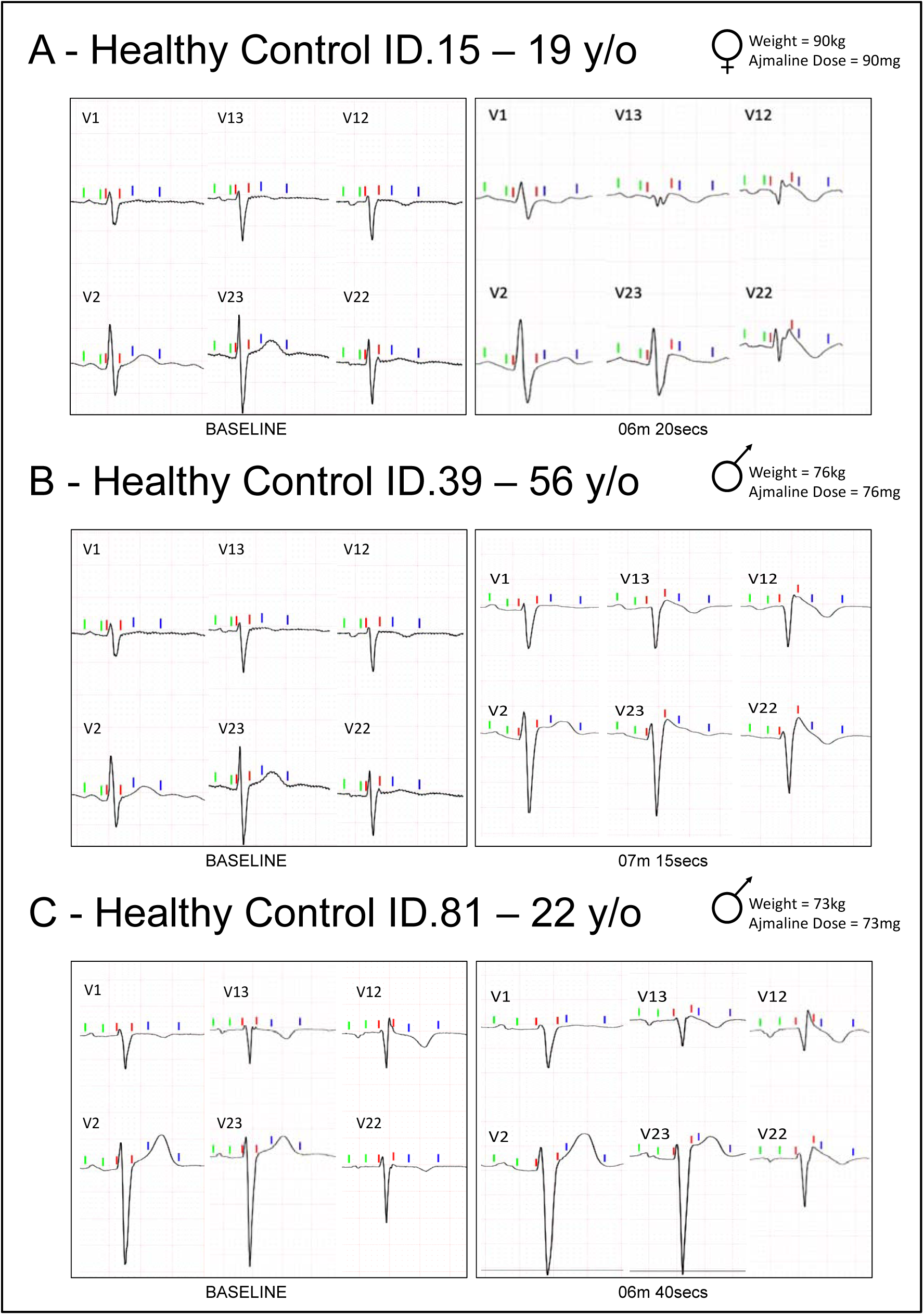
ECGs for the three healthy controls subjects in which the DI-T1BP was observed during ajmaline provocation.

The 100 HC and 166 Def-BrS subjects were equally and randomly divided into the respective discovery and validation groups. There were no significant demographic or clinical differences between the Def-BrS-Dis and Def-BrS-Val cohorts (Table 3.0). Similarly, there were no demographic differences between the HC-Dis and HC-Val cohorts (Table 4.0).

**Table 3.0.** Comparison between Discovery and Validation Def-BrS cohorts.

|  | Def-BrS-Dis<br>N = 83 | Def-BrS-Val<br>N = 83 | P value |
| --- | --- | --- | --- |
|  | Demographics |  |  |
| Median Age (years) | 42.5 | 42.49 | 0.51 |
| Male Gender | 37 (45%) | 34 (41.%) | 0.64 |
| Caucasian Ethnicity | 64 (77%) | 69 (83%) | 0.33 |
| BrS Proband | 24 (29%) | 27 (33%) | 0.61 |
| Syncope | 27 (33%) | 28 (34%) | 1.00 |
| Baseline T2/3 Brugada pattern | 9 (11%) | 8 (10%) | 0.89 |
|  | Ajmaline Indication |  |  |
| SADS/UCA relative | 20 (24%) | 19 (23%) | 0.82 |
| T1BP in a relative | 50 (60%) | 49 (59%) |  |
| UCA survivor | 4 (5%) | 7 (8%) |  |
|  | Family History Score |  |  |
| 0 | 15 (18%) | 14 (17%) | 0.93 |
| 0.5 | 7 (8%) | 5 (6%) |  |
| 1 | 8 (10%) | 9 (11%) |  |
| 2 | 53 (64%) | 54 (66%) |  |
| Mean (SD) | 1.42 (0.82) | 1.46 (0.80) | 0.74 |
|  | Clinical History Score |  |  |
| 0 | 54 (65%) | 55 (66%) | 0.10 |
| 0.5 | 1 (1%) | 1 (1%) |  |
| 1 | 11 (13%) | 17 (21%) |  |
| 2 | 9 (11%) | 1 (1%) |  |
| 3 | 8 (10%) | 9 (11%) |  |
| Mean (SD) | 64 (1.02) | 0.56 (0.96) | 0.58 |
|  | ECG Score |  |  |
| 0 | 0 (0%) | 0 (0%) | 0.47 |
| 2 | 69 (83%) | 71 (86%) |  |
| 3 | 2 (2%) | 4 (5%) |  |
| 3.5 | 12 (15%) | 8 (10%) |  |
| Mean (SD) | 2.24 (0.54) | 2.19 (0.48) | 0.55 |
|  | Genetic Score |  |  |
| 0.5 | 16 (19%) | 15 (18%) | 0.84 |
| Mean (SD) | 0.10 (0.20) | 0.10 (0.19) | 0.84 |
|  | Ajmaline Provocation |  |  |
| Pre - ajmaline SS (SD) | 2.16 (0.81) | 2.09 (0.75) | 0.59 |
| Final SS (SD) | 4.40 (0.85) | 4.30 (0.63) | 0.33 |
| Time to Peak Drug Effect (SD) | 04m59s (02:19) | 05m05s (01:22) | 0.08 |
| Final dose:Target dose (SD) | 0.77 (0.23) | 0.79 (0.21) | 0.65 |

**Table 4.0.** Comparison between Discovery and Validation HC cohorts. Def-BrS-Dis= Definite Brugada Syndrome discovery group.

|  | **HC-Dis**<br>**N=50** | **HC-Val**<br>**N=50** | **P value** |
| --- | --- | --- | --- |
| **Median Age in years**<br>**(Q1-Q3)** | 22.99<br>(22.91 - 30.38) | 23.26<br>(20.86 - 26.79) | 0.03 |
| **Mean age in years (SD)** | 27.85 (7.45) | 25.83 (8.46) | 0.21 |
| **Male Gender N (%)** | 26 (52%) | 26 (52%) | 0.58 |
| **Weight in Kg (SD)** | 74.98 (13.30) | 75.49 (11.52) | 0.84 |
| **Time to peak effect**<br>**(mm:ss) (SD)** | 05:51 (01:28) | 06:08 (01:10) | 0.30 |
| **Drug-induced Type 1**<br>**Brugada Pattern** | 4% (2/50) | 2% (1/50) | 1.00 |

The independent sample univariable analysis of baseline ECG characteristics demonstrating a statistically significant difference between the discovery groups is highlighted in Table 5.0. Similarly, Table 6.0 highlights the univariable analysis of statistically significant ΔECG characteristics.

**Table 5.0.** Univariable analysis of statistically significant baseline ECG characteristics between the discovery groups. §Net area.

|  | **Def-BrS-Dis**<br>**N=83**<br>**Mean** | **HC-Dis**<br>**N=50**<br>**Mean** | **P Value** | **95% Confidence Interval of the Difference** |
| --- | --- | --- | --- | --- |
| **QRS duration in ms (SD)** | 99.49 (19.53) | 90.56 (9.66) | <0.01 | 3.08 - 14.79 |
| **T wave duration in ms (SD)** | 192.94 (33.73) | 206.44 (25.76) | 0.02 | -24.48 - -2.53 |
| **PR interval in ms (SD)** | 169.18 (27.22) | 151.48 (20.14) | <0.01 | 8.91 - 26.49 |
| **QT interval in ms (SD)** | 390.39 (32.38) | 404.28 (29.37) | 0.01 | -24.97 - -2.81 |
| **P wave duration in ms (SD)** | 113.57 (15.83) | 103.00 (9.89) | <0.01 | 5.64 - 15.49 |
| **S wave duration II in ms (SD)** | 27.45 (20.19) | 18.40 (14.98) | <0.01 | 2.52 - 15.57 |
| **S wave amplitude I in μV (SD)** | -129.16 (157.76) | -191.62 (174.73) | 0.04 | 4.27 - 120.65 |
| **Precordial lead QRS duration in ms (SD)** | 94.00 (18.15) | 85.87 (9.09) | <0.01 | 2.68 - 13.59 |
| **Anterior lead ST Amplitude in μV (SD)** | 73.87 (65.87) | 44.43 (32.34) | <0.01 | 9.70 - 49.18 |
| **Precordial lead T wave duration in ms (SD)** | 159.72 (34.77) | 171.40 (27.76) | 0.05 | -23.13 - -0.23 |
| **§Precordial lead QRS Area in mV/ms (SD)** | -283.15 (339.23) | -392.47 (264.15) | 0.05 | -1.63 - 220.25 |
| **§Precordial lead STj_Tp Area in mV/ms (SD)** | 353.17 (215.65) | 483.14 (244.65) | <0.01 | -210.34 - -49.60 |
| **Baseline Type 2/3 pattern N (%)** | 9 (10.84%) | 0 (0%) | 0.02 | - |

**Table 6.0.** Univariable analysis of statistically significant ΔECG characteristics between the discovery groups. § Net Area.

|  | **Def-BrS-Dis**<br>**N=83** |  | **HC-Dis**<br>**N=50** |  | **P**<br>**Value** | **95%**<br>**Confidence**<br>**Interval of the**<br>**Difference** |
| --- | --- | --- | --- | --- | --- | --- |
|  | **Mean** | **(SD)** | **Mean** | **(SD)** |  |  |
| **QRS duration (ms)** | 36.27 | (19.26) | 26.80 | (12.87) | <0.01 | 3.39 - 15.54 |
| **PR interval (ms)** | 19.45 | (31.80) | 42.08 | (16.48) | <0.01 | 13.03 – 32.23 |
| **P wave duration**<br>**(ms)** | 8.82 | (25.22) | 19.12 | (12.50) | <0.01 | -17.87 - -2.73 |
| **S wave duration lead**<br>**II (ms)** | 26.27 | (21.11) | 16.36 | (15.67) | <0.01 | 3.09 - 16.72 |
| **Anterior ST**<br>**amplitude (µV)** | 134.99 | (114.85) | 26.55 | (27.76) | <0.01 | 52.95 - 123.94 |
| **Precordial lead QRS**<br>**Area (mV/ms)<sup>§</sup>** | 124.43 | (297.36) | -186.80 | (187.76) | <0.01 | 218.52 - 403.94 |

Following a combined multivariable analysis of baseline and ΔECG characteristics significantly in favour of Def-BrS (OR >1.0 and P≤0.05) in the discovery cohort, anterior ST amplitude (Adj. OR. 1.027 [95% Confidence Interval 1.005 – 1.046], P= 0.014) and QRS duration (Adj. OR. 1.096 [1.010 – 1.190], P=0.028) were independent baseline predictors of Def-BrS. The ΔAnterior ST amplitude (Adj.OR.1.027 [1.012 – 1.043]), P<0.001, ΔQRS area (Adj. OR. 1.008 [1.004 – 1.013], P<0.001 and ΔQRS duration (Adj. OR. 1.127 [1.050 – 1.209], P<0.001) were SCBP associated ECG characteristics independently predictive of Def-BrS (Table 7.0).

**Table 7.0.** Combined multivariable analysis of baseline & ΔECG characteristics between the discovery groups. *Per 1 unit increase in variable for quantitative traits. ^§^Net area.

|  | **Adjusted Odds Ratio\*** | **P Value** | **95% Confidence Interval** |
| --- | --- | --- | --- |
| **Baseline Type 2/3 Brugada Pattern** | 1x10<sup>9</sup> | 0.999 | 000 - . |
| **Baseline QRS duration (ms)** | 1.096 | 0.028 | 1.010 - 1.190 |
| **Baseline P wave duration (ms)** | 1.054 | 0.910 | 0.992 - 1.120 |
| **Baseline S wave duration lead II (ms)** | 0.969 | 0.179 | 0.925 - 1.015 |
| **Baseline Anterior lead ST Amplitude (uV)** | 1.026 | 0.014 | 1.005 - 1.046 |
| **Baseline precordial QRS area (mV/ms) <sup>§</sup>** | 1.000 | 0.916 | 0.997 – 1.003 |
| **ΔQRS duration (ms)** | 1.127 | <0.001 | 1.050 – 1.209 |
| **ΔS wave duration lead II (ms)** | 0.967 | 0.232 | 0.916 – 1.021 |
| **ΔAnterior lead ST Amplitude (uV)** | 1.027 | <0.001 | 1.012 – 1.043 |
| **ΔPrecordial lead QRS area (mV/ms) <sup>§</sup>** | 1.008 | <0.001 | 1.004 – 1.013 |

The adjusted odds ratios per 10-unit increment for the final ECG characteristics in the Def-BrS diagnostic ECG model are highlighted in Figure 3.0. The final ECG model was 92.8% sensitive and 90.0% specific, with an overall accuracy of 91.7% for Def-BrS and predicted the correct group membership in the 2 HC in the discovery cohort with a DI-T1BP. With an area under the receiver operator curve (AUROC) of 0.95 [0.912 – 0.989], P<0.001, the combined model was an outstanding discriminator for the diagnosis of Def-BrS. The optimal predictive probability from the derived ECG model was 0.617 (with 0.00= healthy control and 1.00= definite BrS syndrome) which corresponded with a sensitivity of 92.8% and specificity 96.0% (Figure 4.0).

**Figure 3.0.**
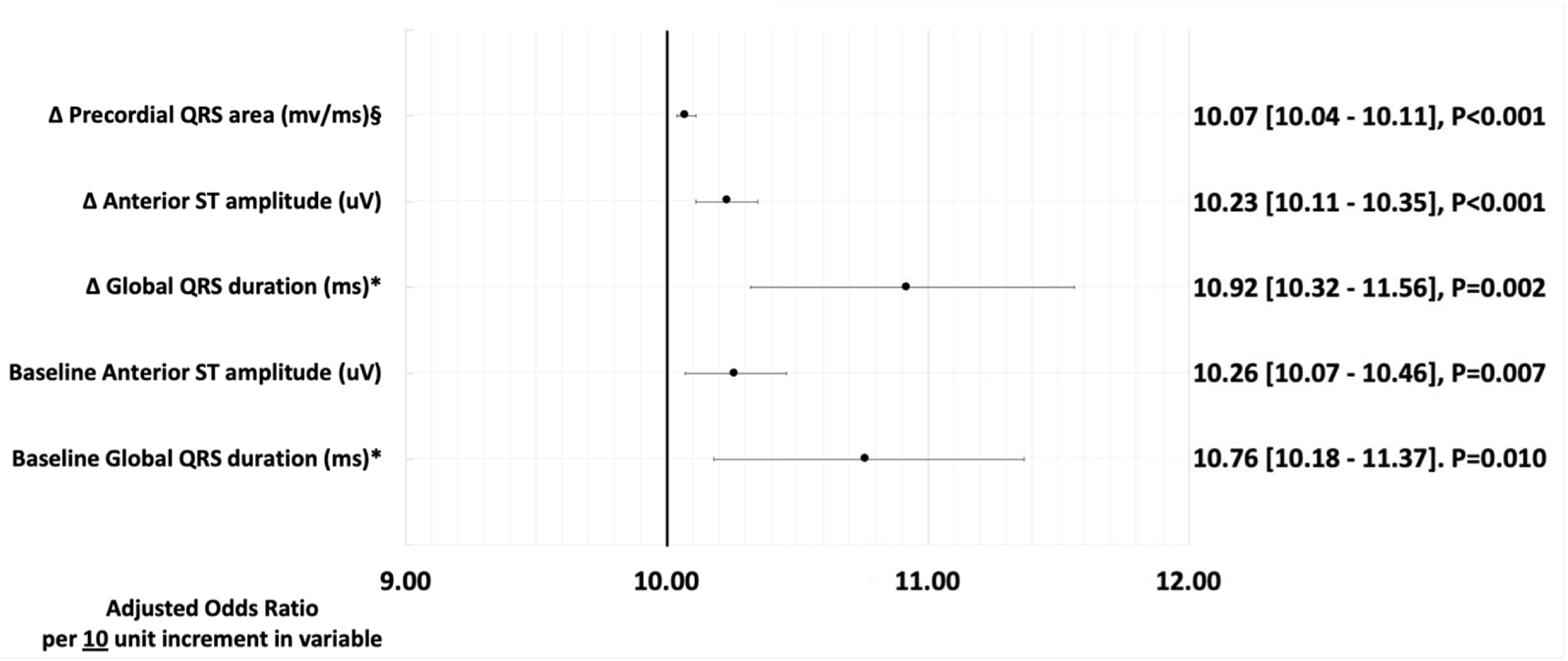
Forrest Plot of final ECG characteristics and corresponding adjusted odds ratios per 10unit increment in value. ^§^Net area.

**Figure 4.0.**
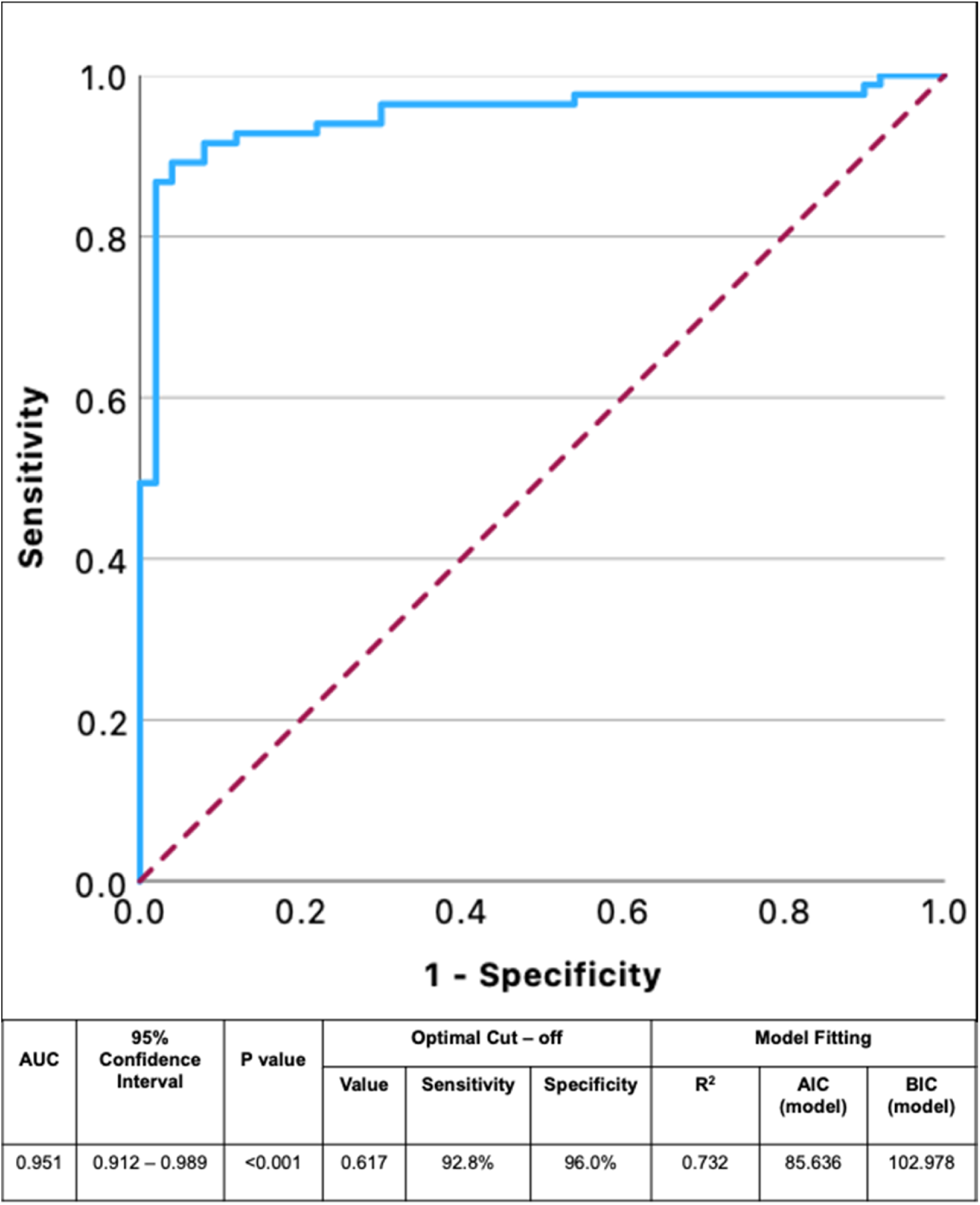
ROC curve for the Def-BrS predictive ECG discovery model. AUC=Area under the curve.

In the validation group, a similar performance was observed with an AUROC = 0.97 [0.948 – 0.998], P<0.001. Similar levels of sensitivity and specificity were observed 97.6% and 90.0% respectively with an overall accuracy of 94.7%. The optimal predictive probability was 0.502 which corresponded to a sensitivity of 97.6% and specificity of 99%. Again, the model predicted the correct group membership in the 1 HC in the validation group with a DI-T1BP

The AIC of the intercept only was 178.103 and 85.636 with the final model suggesting a good fit. Similarly, the BIC of the intercept only was 180.993 and 102.978 with the final model. The *R*^2^ for the overall model was 0.732.

## DISCUSSION

We report the first systematic assessment of diagnostic SCBP with ajmaline in a large cohort of asymptomatic healthy Caucasian subjects without i) a family history of sudden or unexplained death, ii) ECG or structural abnormalities. We identified that the DI-T1BP may be observed in 3% of healthy individuals. In addition, baseline anterior ST segment amplitude, baseline global QRS duration, Δ precordial QRS area, Δ anterior ST amplitude and Δ global QRS duration were independently associated with a diagnosis of *definite* concealed BrS. In combination, these ECG characteristics were able to accurately distinguish between the healthy and those with Def-BrS with a high level of specificity and sensitivity. Therefore, these quantitative characteristics can enhance diagnostic certainty for BrS.

### Comparison to previously reported yields

In clinical cohorts, the yield of the DI-T1BP with ajmaline is variable; in relatives of SADS victims a yield of 14-15% has been observed ^(1, 8)^. However, in survivors of UCA, a yield of 20-22% has been reported ^(8, 9)^. Hasdemir *et al* observed a yield of the DI-T1BP with ajmaline of 27.1% in cohort of Turkish patients with a diagnosis of atrioventricular nodal re-entrant tachycardia (AVNRT) but without a baseline ECG suspicious for BrS ^(4)^. Of note, in those with AVNRT and a DI-T1BP who underwent genetic testing, a variant in a gene related to sodium channel current was identified in 76.5%. However, pathogenicity of these variants was uncertain and baseline conduction intervals (PR, HV and QRS) were normal, rendering these variants unlikely to be clinically relevant. Interestingly, in the study by Hasdemir *et al*, a DI-T1BP was observed in 4.5% of the control subjects who were unrelated and free from any atrial and ventricular arrhythmia. In contrast to our study, neither family history nor a history of syncope were used to screen controls. The false positive rate of SCBP with ajmaline is therefore likely to be between 3.0-4.5% in individuals without cardiovascular symptoms. However, the false positive rate may be higher in those with a history of palpitations suggestive of AVNRT, and clinicians must be aware of this prior to SCBP.

### Baseline ECG characteristics

A T2/3BP was observed at baseline in 10% of patients with Def-BrS was not seen in any of the HCs, and whilst the odds ratio in the multivariable logistic regression analysis was exponential it was not a statistically significant independent predictor of the diagnosis of Def-BrS in the discovery cohort (OR 1×10^9^, P=0.99). The true significance of a baseline T2/3BP is therefore out of the scope of this study and must be assessed in a larger cohort of clinical patients, including those with low and intermediate diagnostic Shanghai scores.

In the high lead configuration right ventricular conduction delay or an incomplete right bundle branch pattern can be observed in healthy subjects ^(10)^. Several ECG features have been proposed to aid in the differentiation between a benign r’ and a T2/3BP at baseline(11-14), including an assessment of the ß angle, which in clinical practice might be challenging to measure. In the combined predictive ECG model, we observed that baseline ST elevation in the precordial leads and QRS duration were independent predictors of the diagnosis of definite Brugada syndrome, with an adjusted OR per 10µV of 10.26 (95% CI 10.07 – 10.46), P= 0.007 and 10.76 (95% CI 10.18 −11.37), P= 0.010. These parameters can be measured routinely with a consistent degree of reproducibility in clinical practice.

### Ajmaline Provoked ECG characteristics

The change in global QRS duration with ajmaline was significantly greater in the Def-BrS-Dis group compared to the HC-Dis group. However, ΔPR interval and ΔP wave duration were significantly greater in the healthy controls. In our study, as with most SCBP protocols, intravenous administration stopped once the DI-T1BP was observed. As a result, the mean actual dose given in the Def-BrS-Dis cohort was 78% of the target dose, while the mean actual dose given in the HC-Dis subjects was 100% of the target dose (P<0.01). This might suggest that mechanistically, the observed differences in the ΔPR interval and ΔP wave duration may be dose dependent, whilst the effects on QRS duration and S wave duration in lead II are disease dependent.

SCBP resulted in a significant increase in ST segment amplitude in the anterior leads in the Def-BrS-Dis group compared to the HC-Dis group. The mean baseline anterior ST amplitude in the Def-BrS-Dis group was 73.87*µ*V (±65.87), and at peak drug effect was 208.86*µ*V, above the threshold required for a DI-T1BP (>0.2mV). This would suggest that in a proportion of the Def-BrS-Dis group, the DI-T1BP was observed prior to peak drug effect, which in the Def-BrS-Dis group was 04m59s (±02m19s).This is suggestive of a lesser capacity of the RVOT in Def-BrS to withstand the insult of SCBP and therefore less RVOT conduction reserve ^(7)^. However, the numbers of HCs with a DI-T1BP were too small to conder this a reliable co-variate.

### Predictive ECG model

In a combined model, the baseline and ΔECG characteristics that were independently associated with the Def-BrS group (Figure 3.0) predicted group membership with a high level of specificity and sensitivity in the discovery cohort, AUC 0.95, P<0.01. The optimal cut-off value for Def-BrS had 92.8% sensitivity and 96.0% specificity. In the validation cohort a similarly high level of accuracy was observed. These findings suggest that a quantitative refinement on the ECG score will enhance the diagnostic power of ajmaline provocation from the binary assessment that currently exists, enabling the identification of those with a firm diagnosis even in the absence of clinical or genetic characteristics that were not incorporated in this study.

While an external validation of the predictive model in a similar clinical cohort of patients with a DI-T1BP and SS ≥3.5 is desirable, an assessment of the model’s performance in a cohort of patients with a DI-T1BP with low or intermediate diagnostic certainty is necessary to determine whether a diagnosis could be “*upgraded*” or “*downgraded*” based on the quantitative ECG score. Finally, an assessment of the association between the ECG model and clinical variables, such as family history, symptoms and genetic information may determine whether these findings can become a ‘gold standard’ for the diagnosis of concealed BrS.

### Clinical impact of this study

For the first time in a large cohort of systematically screened Caucasian ‘super-healthy’ subjects, we have determined that a DI-T1BP can be observed in 3%. In clinical practice, clinicians will be guided by this observation to better inform their patients when considering whether to undertake SCBP with ajmaline. The ECG traits in favour of the diagnosis of definite BrS are readily measurable but the accuracy is greatly enhanced by the ability to acquire the ECG digitally and perform a subsequent automated analysis. This should be considered the ideal method for ECG acquisition when performing SCBP.

### Limitations

Our study used ajmaline, but whether these findings can be extrapolated to other sodium channel blockers is uncertain and should be addressed using a comparative and collaborative approach. Eighty six percent (86%) of the discovery cohort in the study were Caucasian and thus whether the model can be applied to other ethnicities is also uncertain.

## CONCLUSIONS

By leveraging a novel clinical trial of ajmaline in the healthy, we have identified disease-specific ECG changes that are present at baseline and with SCBP that may enhance the diagnostic certainty of a DI-T1BP, and thus lay the groundwork for a gold standard for the diagnosis of concealed Brugada syndrome. We also demonstrated that changes in healthy subjects are observed with ajmaline, including the development of a DI-T1BP in 3%, that may indicate physiological variation in RVOT conduction reserve that is likely to be more impaired in true BrS.

## SUPPLEMENTARY PUBLICATION MATERIAL

**I – ONLINE QUESTIONNAIRE**

**I – INCLUSION/EXCLUSION CRITERIA**

**I – HIGH RIGHT PRECORDIAL ECG PLACEMENT**

**I – ECG FIGURES**

**I – PAIRED ANALYSIS**

## Data Availability

All data is available on request

## Acknowledgements

Dr Velislav Batchvarov MD (posthumous), Cardiology Clinical Academic Group, St. George’s University Hospitals NHS Foundation Trust, London, UK and Institute of Molecular and Clinical Sciences, St George’s University of London, London, UK,

